# MyoVision-US: an Artificial Intelligence-Powered Software for Automated Analysis of Skeletal Muscle Ultrasonography

**DOI:** 10.1101/2024.04.26.24306153

**Authors:** Zoe Calulo Rivera, Felipe González-Seguel, Arimitsu Horikawa-Strakovsky, Catherine Granger, Aarti Sarwal, Sanjay Dhar, George Ntoumenopoulos, Jin Chen, V. K. Cody Bumgardner, Selina M. Parry, Kirby P. Mayer, Yuan Wen

**Author notes:** Denotes co-first author with authors contributing equally (Zoe Calulo Rivera, Felipe González-Seguel, Arimitsu Horikawa-Strakovsky). Denotes co-senior author with authors contributing equally (Selina M. Parry, Kirby P. Mayer, Yuan Wen).

## Abstract

**Introduction/Aims:** Muscle ultrasound has high utility in clinical practice and research; however, the main challenges are the training and time required for manual analysis to achieve objective quantification of morphometry. This study aimed to develop and validate a software tool powered by artificial intelligence (AI) by measuring its consistency and predictability of expert manual analysis quantifying lower limb muscle ultrasound images across healthy, acute, and chronic illness subjects.

**Methods:** Quadriceps complex (QC [rectus femoris and vastus intermedius]) and tibialis anterior (TA) muscle ultrasound images of healthy, intensive care unit, and/or lung cancer subjects were captured with portable devices. Automated analyses of muscle morphometry were performed using a custom-built deep-learning model (MyoVision-US), while manual analyses were performed by experts. Consistency between manual and automated analyses was determined using intraclass correlation coefficients (ICC), while predictability of MyoVision -US was calculated using adjusted linear regression (adj.R^2^).

**Results:** Manual analysis took approximately 24 hours to analyze all 180 images, while MyoVision - US took 247 seconds, saving roughly 99.8%. Consistency between the manual and automated analyses by ICC was good to excellent for all QC (ICC:0.85–0.99) and TA (ICC:0.93–0.99) measurements, even for critically ill (ICC:0.91–0.98) and lung cancer (ICC:0.85–0.99) images. The predictability of MyoVision-US was moderate to strong for QC (adj.R^2^:0.56–0.94) and TA parameters (adj.R^2^:0.81–0.97).

**Discussion:** The application of AI automating lower limb muscle ultrasound analyses showed excellent consistency and strong predictability compared with human analysis. Future work needs to explore AI-powered models for the evaluation of other skeletal muscle groups.

## INTRODUCTION

Peripheral muscle dysfunction can encompass reductions in muscle mass, strength, endurance, and fatigability.^1^ Muscle ultrasound imaging enables non-invasive bedside evaluation of skeletal muscle quantity (i.e., muscle thickness, anatomical cross-sectional area [CSA]), quality (echointensity [EI]) and architectural properties (i.e., pennation angle, fascicle length, contraction potential).^1,2^ It has become increasingly popular as an assessment tool to evaluate longitudinal changes in muscle characteristics and to evaluate intervention efficacy in relation to exercise, nutrition and/or pharmacological interventions targeting muscle in healthy and diverse patient populations.^1,3–6^ Current literature is focused on muscle ultrasound evaluation of the quadriceps muscle group due to its size and importance in physical function, with rectus femoris (RF) muscle thickness considered to be a reliable reflection of muscle strength.^7–9^ Muscle ultrasound imaging has demonstrated good to excellent inter- and intra-rater reliability and strong criterion validity compared to histochemistry analyses on muscle biopsy in older adults,^10^ individuals with myopathic disease,^11^ and intensive care unit (ICU) survivors 6-12 months after discharge.^12^

The benefits of muscle ultrasound support the utilization of this tool in clinical and research practice. However, the ability of parameters defined from muscle ultrasound to predict meaningful outcomes is equivocal.^13,14^ Limited predictive validity may partially be explained by sonographers using a subjective scale at the bedside (ie. the Heckmatt approach for qualitative evaluation of EI)^15^ or the need for a trained expert to manually analyze ultrasound images for objective parameters.^16–18^ The objective analysis of muscle parameters from ultrasound images is manual operator-dependent, requiring sustained human engagement that is time and labor-intensive, increasing the potential for human biases. Technology provides an avenue to enhance image analyses by developing programs for automation with artificial learning.^19^ Computational programs capable of mimicking human decision-making behavior are broadly known as artificial intelligence (AI). Machine learning is a branch of AI and involves the process of building algorithms to learn patterns or rules based on data.^20^ Automated methods of analyzing ultrasound images have been investigated to address the limitations created by manual data interpretation. In the last decade, the use of AI and machine learning in medical research, particularly medical image analysis, has grown in popularity, as these models have been shown to improve efficiency and lower the error rate of clinical tasks.^20^ Recent studies by Cronin et al. and Katakis et al. have shown that machine and deep learning can improve existing automated systems and produce comparable results to manual analysis, indicating that these methods are both acceptable and reliable.^21–23^ In 2017, Caresio et al. developed and validated a fully automatic method, named MUSA (Muscle UltraSound Analysis), for measuring muscle thickness on longitudinal ultrasound images.^24^ Despite its accuracy, this method was limited to the analysis of muscle thickness in healthy individuals. This highlights the need to develop automated methods of analyzing a wider range of muscle ultrasound parameters for use in healthy and diseased populations.

MyoVision is a freely available automated image analysis software that was originally developed for the quantification of muscle cross-sections from biopsies in animal models and human studies.^25,26^ It is a reliable and accurate program that automates the analyses of immunohistochemistry, which has been utilized worldwide and heavily cited since its original release in 2018.^25,26^ The success of MyoVision and the need for automated software for ultrasonography led to the development of MyoVision -US that provides comprehensive automated quantification of lower limb muscle thickness, CSA, and EI without human supervision. Therefore, this study aimed to develop and validate the AI-powered MyoVision-US by measuring its consistency and predictability of expert manual analysis of muscle thickness, CSA, and EI of ultrasound images across healthy, acute, and chronic illness cohorts.

## METHODS

### Study Design

A prospective study for the development and validation of MyoVision -US was conducted in accordance with prior work,^25,26^ and data was compared to manual analysis of muscle ultrasound images. The reporting of clinimetric properties was performed in accordance of the COnsensus-based Standards for the Selection of Health Measurement Instruments (COSMIN) guidelines.^27^ This study received approval from the Nonmedical Institutional Review Board (IRB #75185) and the Data Use Agreement of the University of Kentucky for image sharing across sites.

### Musculoskeletal Ultrasound Image Dataset

A diverse dataset of muscle ultrasound images was developed from active (ACTRN12618001733268; ACTRN12617001283369; NCT03141762) and completed studies.^4,7,17,28–31^ Prior to dataset transfer all personal or patient identifiers were removed from images or data files. A test set was curated to assess the performance of the automated MyoVision software. A blinded researcher randomly selected 90 ultrasound images of the quadriceps complex (QC) including 30 from individuals with critical illness in the ICU or during the recovery phase, 30 from individuals with lung cancer with outpatient scans, and 30 volunteer healthy adult individuals; all images were obtained from adults (≥ 18 years of age). Additionally, 90 tibialis anterior (TA) ultrasound images were also randomly extracted including 60 selected from the same individuals in the ICU and 30 selected from the same volunteer healthy individuals. The three adult populations were selected to provide a spectrum of muscle health to enhance generalizability during the validation ranging from healthy, acute and chronic illness states.

### Image Quantification

Manual image analysis was performed by three independent physiotherapists (SMP, KPM, and FGS) with at least seven years of muscle ultrasound practical experience^7,12,13,17,18,28,30–34^ using freely available NIH Image J software.^35^ SMP has 14 years of theoretical and practical ultrasound imaging expertise, including training of other clinicians in the use of muscle ultrasound in clinical and research practice internationally at conferences and workshops. KPM has over 9 years of experience utilizing muscle ultrasound in research and practice, including leading educational sessions in the United States. FGS has over 8 years of experience utilizing muscle ultrasound in research and practice and has led Chilean educational courses on muscle ultrasound. All have published papers in both clinical and educational contexts related to muscle ultrasound imaging.^7,12,13,17,18,28,30–34^

For manual analyses, all muscle ultrasound parameters (muscle thickness, anatomical CSA, and EI) were analyzed in triplicate where the mean of the three values used in the final analyses. All muscle thickness parameters were measured in centimeters. Rectus femoris thickness was measured between the two transverse fascial planes of the muscle belly utilizing the center of the femur for consistency. Vastus intermedius (VI) was measured between the uppermost part of the femur bone to the superficial fascia of the VI muscle utilizing the center of the femur for consistency. The thickness of QC was measured between the uppermost part of the bone echo of the femur and the superficial fascia and includes both the RF and VI muscles. Tibialis anterior was measured from the superficial fascia to the uppermost part of the bone echo of the tibia. Cross-sectional area was measured in square centimeters using the free-form mode to trace the inside border of the epimysium for both RF and TA muscles. Echointensity was quantified based on the grey scale of the pixels of the image ranging from 0 (black) to 255 (white) for RF and TA muscles using two approaches: 1) the mean and standard deviation of the grayscale values were calculated using the square trace method, utilizing a bespoke distance box approach as previous described^30,36^ and 2) greyscale of the pixels calculated using the free-form trace outline method described to determine CSA.

### MyoVision-US software implementation

As shown in **Figure 1**, the MyoVision-US workflow consists of three steps. The first step is the prediction by a DeepLabV3^37^ model, followed by the second step of post-processing, and the last step involves the calculation of muscle image parameters.

**Figure 1.**
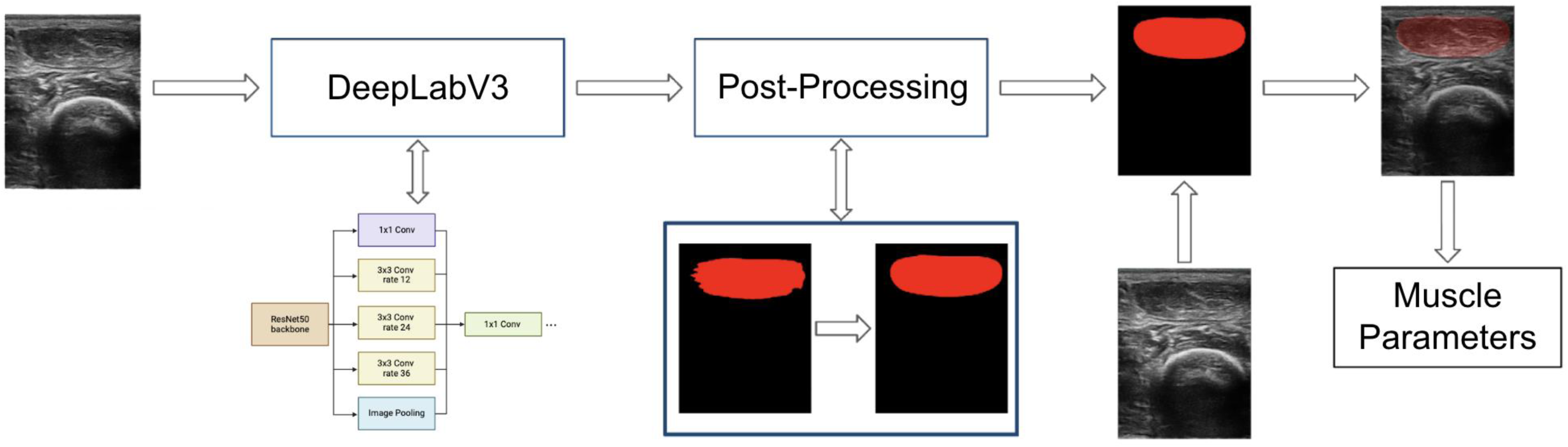
Diagram of MyoVision-US workflow, with 3 stages: DeepLabV3 model, post-processing, and value calculation. The analysis of the rectus femoris muscle in an ultrasound image of the quadriceps complex is specifically shown.

**Step 1:** 2 DeepLabV3 models with a ResNet50^38^ backbone were used, one for QC ultrasound images and one for TA ultrasound images. DeepLabV3 is a deep, fully convolutional neural network architecture for semantic segmentation, which utilizes a backbone to initially identify and extract useful features from the input image and cascades of atrous convolutions to recognize and segment objects at multiple scales. The ResNet50 model is a residual network architecture that introduced skip connections and is commonly used for computer vision and image classification tasks. The dataset used to train the models consists of 94 QC images and 243 TA images representing a mixed group of ICU, lung cancer, and healthy individuals (this set is separate from the validation dataset described in previous sections). These images were then labeled by experts with polygon traces of the maximum visible CSA of the RF, VI, and femur on QC images and the TA on TA images. Images and labels were normalized to stabilize training and resized to 512x512 pixels. Spatial and color data augmentation was also used and was performed using Albumentations^39^, a python library used for image augmentation in computer vision tasks. The ResNet50 backbone was initialized with pretrained ImageNet^40^ weights. The whole model was then trained on either the QC or TA datasets for 40 epochs using a cosine annealing learning rate scheduler^41^ with linear warm up, a maximum learning rate of 5e-4 and AdamW^42^ with a weight decay of 1e-6. The models were evaluated using the Dice coefficient^43^, a measure of similarity between predicted and ground truth labels ranging from 0 being no spatial agreement to 1 being complete agreement (in this case, Dice coefficient is used to compare model predicted and expert labelled maximum visible CSA of muscles and bone). The trained DeepLabV3 models of QC and TA achieved Dice coefficients of 0.81±0.08, and 0.93±0.01 (mean ± standard deviation). The best performing QC and TA models among the 5 trained for each was then chosen for use in the MyoVision -US software.

**Step 2:** A post-processing scheme was used to minimize noise and artifacts in the model’s prediction similar to Katakis et al ^23^. Post-processing starts with the extraction of contours (boundaries of all distinct polygons) in the output prediction (mask) from the model. Then, the contour with the largest internal area was kept and morphological operations, opening and closing, were performed to remove noise by filling in gaps and removing protrusions. In addition, a cubic spline was fit along the contour of the mask to further smooth it.

**Step 3:** After post-processing, calculations of specific values were made. Muscle thickness was calculated by finding the number of pixels predicted to be the target muscle at either the midpoint of the predicted femur mask or the midpoint of the predicted mask of the target muscle (i.e. RF or TA). QC thickness was found by finding the distance, in pixels, at the midpoint of the predicted femur mask between the top of the predicted mask of the femur and the top of the predicted mask of the RF. Cross-sectional area was calculated by using the number of pixels in the predicted mask of the target muscle. Area and thickness measurements were converted to centimeters using the pixel scale for each image calculated from the corresponding depth parameter. Mean and standard deviation of EI was calculated by averaging or finding the standard deviation of the grayscale value of all pixels within the predicted mask of the target muscle.

### Statistical Analysis

Descriptive statistics, histogram plots, and Shapiro-Wilk were performed for all muscle parameters (thickness, anatomical CSA, EI) for both QC and TA muscles. Descriptive statistics are provided for each group (healthy, ICU, and lung cancer) for generalizability. Intraclass correlation coefficients (ICC) were calculated based on the average of measures comparing the software to the manual analysis (95% confidence interval). A two-way random effects test of the average measures was used to examine interrater consistency. The ICC values are scaled from 0 to 1, where an ICC between 0.75 and 0.90 is considered to be good and higher than 0.90 is considered to be excellent reliability.^44^ The correlation between MyoVision -US and manual analysis was reported with Pearson correlation coefficient. To determine the predictability of MyoVision-US (independent variable) and the manual analysis (dependent variable), separate and adjusted linear regression analyses were performed for each muscle parameter. Standard error of measurement (SEM) was reported to estimate the linear regression. The data were analyzed using IBM SPSS Statistics (version 29).

## RESULTS

Ninety QC and 90 TA images were randomly selected to be analyzed by MyoVision-US and compared to manual analyses performed by expert physiotherapists with ultrasound expertise. Manual analysis took experts collectively approximately 24 hours to analyze all 180 images with each image requiring roughly 8 minutes. MyoVision-US took 247 seconds to analyze all 180 images (163 seconds for QC images and 84 seconds for TA), saving roughly 99.8% of the time used for manual analysis. Three of the 90 QC images (3.3%) and five of the 90 TA images (3.4%) were not analyzed in manual analysis due to poor image quality in the ICU group related to marked adiposity or edema, otherwise, there were no missing data. Descriptive statistics for the ultrasound images analyzed by MyoVision-US are provided in **Table 1**. Representative images for RF (**Figure 2A**) and TA (**Figure 2B**) demonstrate the software’s recognition of the muscles of interest for both healthy and ICU patients. MyoVision automated analyses of muscle thickness, CSA, and EI indicate that ICU and/or lung cancer was associated with lower values for RF (**Figure 2C**) and TA (**Figure 2D**) compared to healthy individuals. Direct comparisons between manual and automated analysis of RF (**Figure 2E**) and TA (**Figure 2F**) demonstrate high degrees of consistency for thickness, CSA, and EI.

**Figure 2.**
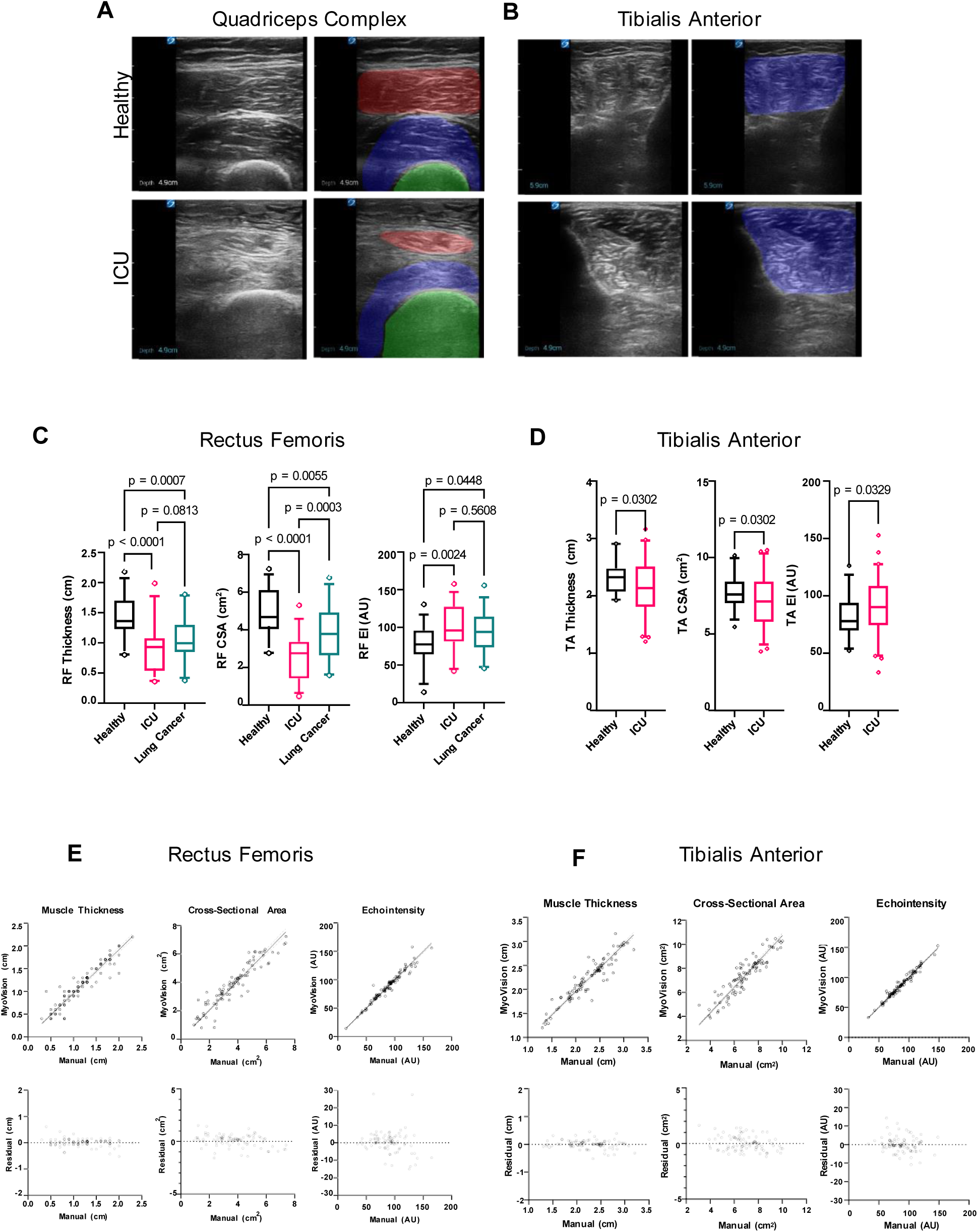
Comparison of automated analysis by MyoVision -US and manual analysis of ultrasound images of quadriceps complex and tibialis anterior. **A-B.** Ultrasound images with and without the free-form trace labeled shadow from MyoVision -US analysis of ultrasound images of the quadriceps complex and tibialis anterior from healthy individuals and patients in the intensive care unit. In ultrasound images of the quadriceps complex, the rectus femoris (red), vastus intermedius (blue), and femur (green) are shown, and in ultrasound images of the tibialis anterior, the tibialis anterior (blue) is shown. **C-D.** Boxplots of rectus femoris muscle thickness, cross-sectional area, and echointensity values of ultrasound images from healthy individuals (black), patients in the intensive care unit (magenta), and patients with lung cancer (green). **E-F.** Scatter plot with regression line plotting values for automated analysis by MyoVision -US against manual analysis of ultrasound images for muscle thickness, cross-sectional area, and echointensity of rectus femoris (left) and tibialis anterior (right). All correlations had p values <0.0001. ICU = intensive care unit, RF = rectus femoris, TA = tibialis anterior, CSA = anatomical cross-sectional area, cm = centimeters

**Table 1.**
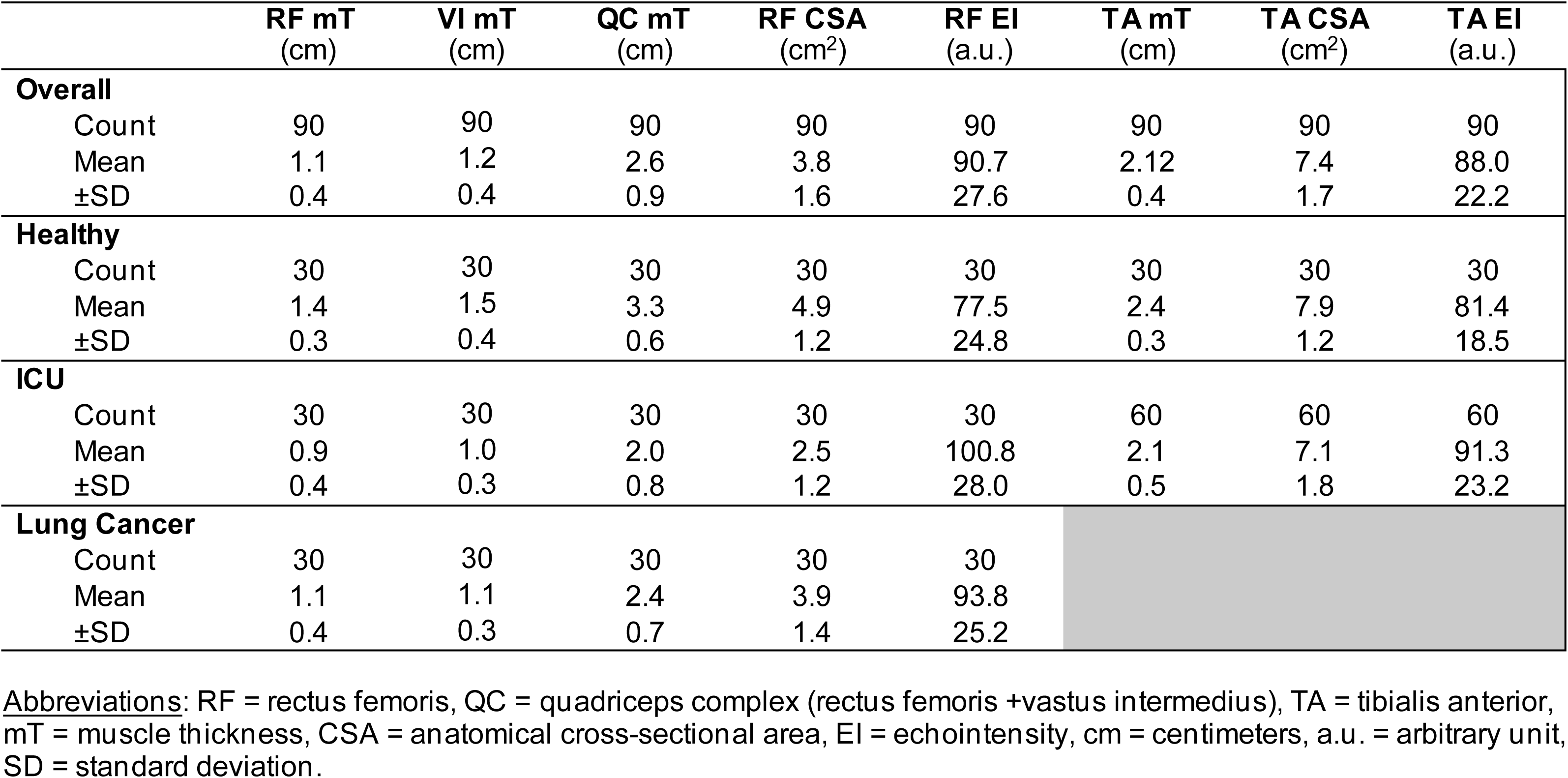
Descriptive statistics of muscle parameters.

The consistency between MyoVision-US and manual analyses for QC images, including examinations of VI and RF for the entire cohort (n = 87), was excellent with ICC ranging from 0.92–0.99 (**Table 2**). Values were not attenuated when examining each group separately, showing good to excellent ICC values for ICU (ICC=0.91–0.98) and lung cancer (ICC=0.85–0.99) images. MyoVision-US demonstrates excellent predictability of the manual analysis with adjusted R^2^ values ranging from 0.76–0.94 in the images of the entire cohort. These values were not considerably lower for ICU (adjusted R^2^=0.68–0.91) and lung cancer (adjusted R^2^=0.55–0.99) images.

**Table 2.**
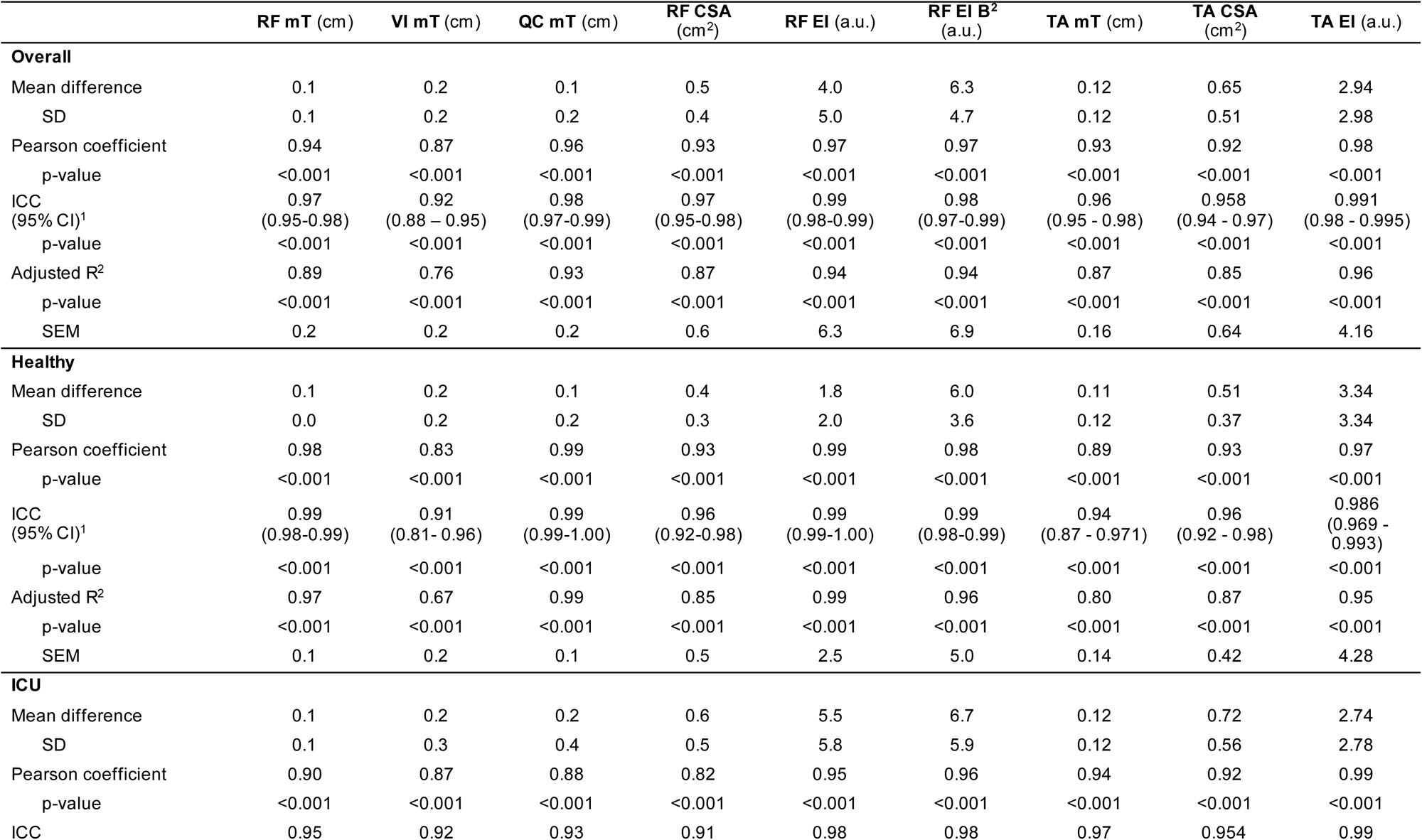
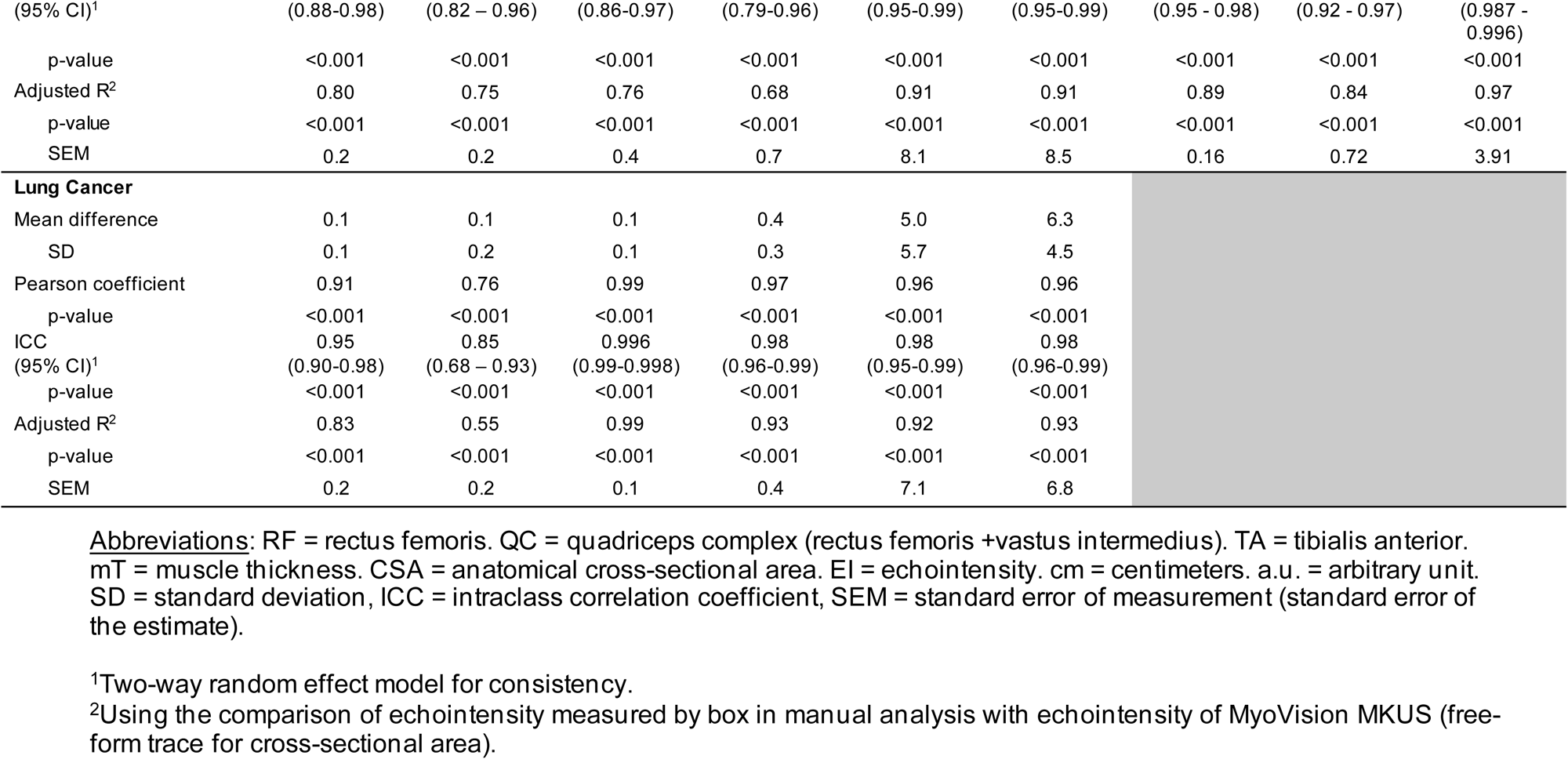
Consistency and predictability of MyoVision-US compared with manual ultrasound analyses.

The consistency between MyoVision-US and manual analyses for TA images of the entire cohort (n = 85) was excellent with ICC ranging from 0.96–0.99 (**Table 2**). Values were not reduced when examining each sub-cohort separately, showing excellent ICC values for ICU (ICC=0.96–0.99) and healthy (ICC=0.94–0.99) individuals. MyoVision-US demonstrates excellent predictability of the manual analysis with adjusted R^2^ values ranging from 0.85–0.96 in the images of the entire cohort. These values were not considerably attenuated for ICU (adj. R^2^=0.84–0.97), where the predictability of MyoVision-US for EI was slightly higher for ICU images (adj. R^2^=0.97) compared to healthy (adj. R^2^=0.95) images.

Regardless of muscle and population group, the predictability by muscle parameter was excellent for EI (adj. R^2^: 0.94–0.96), good to excellent for thickness (adj R^2^=0.76–0.99), and moderate to good for CSA (adj R^2^=0.85–0.87) values (**Table 2**). To facilitate improved utilization of the automated analysis, we developed a simple user interface that allows the upload of an ultrasound image to be analyzed (**Figure 3**). On the left-hand side, the original image can be seen with the accompanying segmentation by the AI model on the right-hand side. We demonstrate the multiclass segmentation capabilities of the models under development and include all anatomical features within the same image, such as the skin, adipose tissue, muscles, and bone. The model and software are available online for demonstration purposes through https://huggingface.co/spaces/ari10/MyoVision-US_Demo.

**Figure 3.**
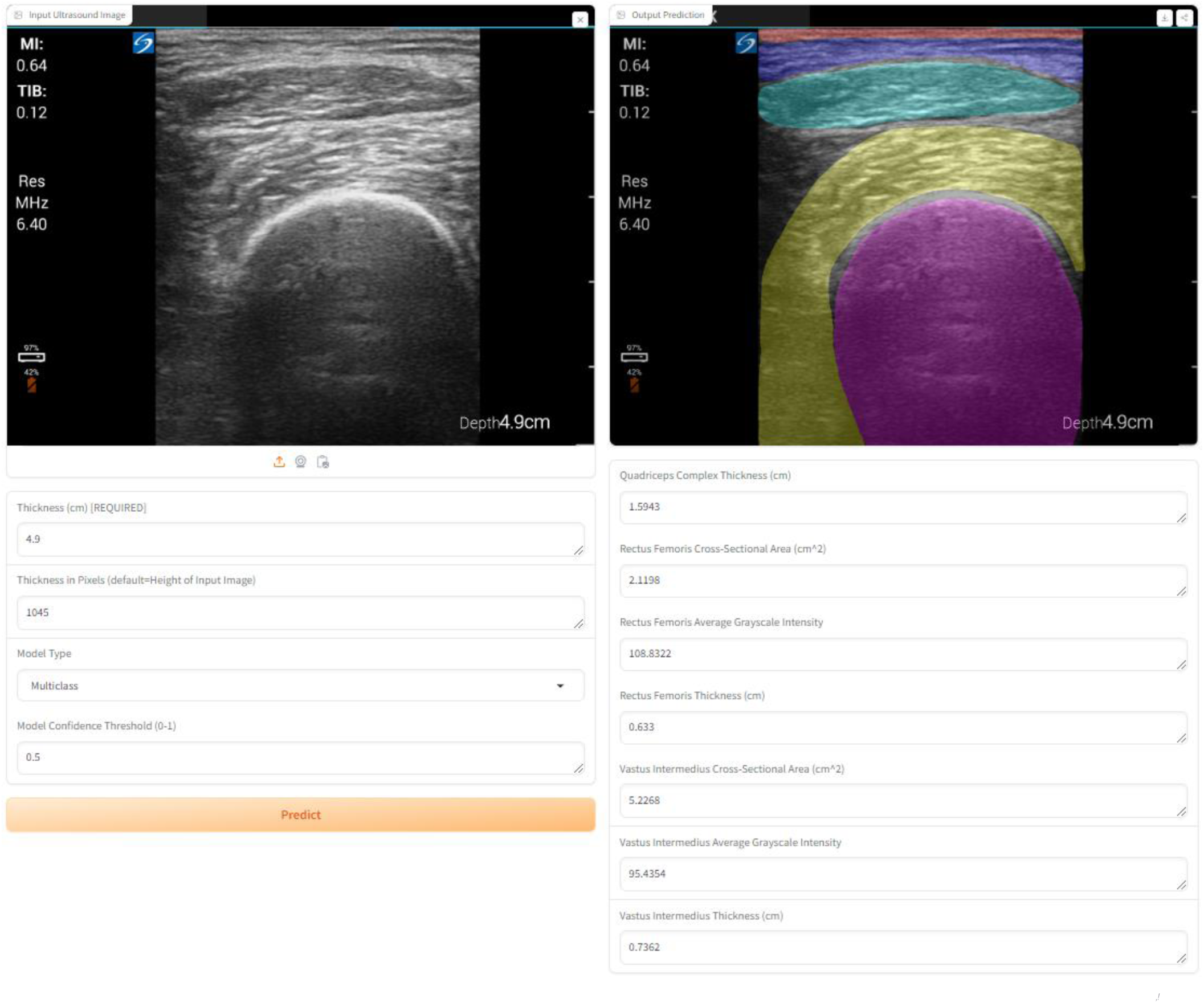
MyoVision-US graphical user interface hosted on HuggingFace, a commonly used platform for building, training, and deploying machine learning models, with results of multiclass analysis and measurements shown. Skin (red), adipose tissue (blue), rectus femoris (yellow), vastus intermedius (aqua), and femur (pink) are segmented and shown.

## DISCUSSION

In this study, MyoVision-US was developed as an AI-driven software for muscle ultrasound image analysis, presenting substantial time savings compared to manual analysis by experts. MyoVision-US and manual analysis had moderate to excellent consistency for QC, RF, VI, and TA ultrasound images of healthy individuals as well as patients with acute and chronic diseases. Furthermore, MyoVision-US demonstrated strong predictability when compared directly to the manual analyses by experts, even for challenging images from patients in the ICU and with lung cancer.

The current description of MyoVision-US implementation represents a very superficial application of the newest AI technologies. Additional development will allow for not only multiclass segmentation from the same image (i.e., delineating all anatomical features, including muscle, bone, skin, adipose, and fascia) but also allow for real-time integration into ultrasound devices. AI-powered precision guidance systems will facilitate efficient and accurate ultrasound image acquisition and open the doors to greater utilization of bedside ultrasounds, which is demanding from an operator training perspective. The generalizability of such AI algorithms for ultrasonography is highly promising for multiple organ systems in addition to and in conjunction with muscle applications ^45^.

Using a deep learning image segmentation model known for performance, this study presents a relevant opportunity to improve the analysis of muscle ultrasound images with clinical and research applications. While previous studies have shown excellent reliability for muscle thickness, muscle CSA, muscle EI, fascicle length, and pennation angle measurements in ultrasound images of healthy muscles ^23,24,46,47^, the present study adds findings with excellent inter-rater consistency for CSA, thickness, and EI of QC and TA muscles in patients with acute and chronic illness. This allowed the spectrum of muscle parameters to be increased, covering high and low values of thickness, CSA, and muscle EI. Muscle quality loss in acute and chronic diseases challenges the quantification of muscle parameters necessary for decision making in clinical practice ^7^. In the present study, MyoVision -US demonstrated excellent consistency and predictability compared to expert manual analysis. Despite the low muscle quality of some challenging images where manual analyses failed, it was possible for the software to measure muscle thickness and CSA, which are useful parameters in populations with severe loss of muscle health (i.e., critical illness and lung cancer) ^3^. Therefore, this new software (MyoVision-US) enhances scientific rigor by reducing human bias while simultaneously reducing the time required to analyze muscle ultrasound images.

This study has potential limitations that need to be addressed. First, the sample size could be viewed as a limitation. However, the inclusion of images from multiple sites encompassing images of healthy individuals and patients with critical illness and lung cancer allowed a wide range of muscle parameters to be covered. Second, the findings of this study could only be generalized to adult healthy individuals or adult patients with critical illness and lung cancer. However, studies on populations with similar muscle conditions could also benefit from automating muscle image analysis. Third, although this study did not describe the accuracy result of each iteration during the development of the MyoVision-US, images were randomly selected and blinded for manual/automated analysis showing excellent consistency with manual analysis. Fourth, the findings of this study are limited to QC and TA muscles, therefore, future reports should explore accuracy in other peripheral and respiratory muscles.

The bedside measurement of muscle parameters has been progressively replaced by post-hoc analysis using software such as NIH Image J mainly to systematize parameters of muscle quantity and quality. However, these analyzes take time and are not exempt from human error. Therefore, the findings of this study become relevant for the interpretation of muscle quality and quantity parameters if adequate image acquisition is performed.

In conclusion, the application of AI to automate muscle ultrasound analyses showed improvements in speed with strong consistency and predictability compared with manual human analysis evaluating muscle thickness, CSA, and EI of QC and TA muscles. This study introduces a method to automate the analysis of lower limb muscle ultrasound images, which may be useful for future research and clinical applications in acute and chronic settings.

## Abbreviations

AI: artificial intelligence
CSA: cross-sectional area
EI: echointensity
ICC: Intraclass correlation coefficient
ICU: intensive care unit
QC: quadriceps complex
RF: rectus femoris
TA: tibialis anterior
VI: Vastus intermedius

## Ethical Publication Statement

We confirm that we have read the Journal’s position on issues involved in ethical publication and affirm that this report is consistent with those guidelines.

## Ethics

IRB #75185 medical expedited review approved February 18, 2022.

## Data sharing

Data is available through reasonable request to the corresponding authors.

## Data Availability

Data is available through reasonable request to the corresponding authors.

